# Deep Learning Models for Radiography Body-part Classification and Chest Radiograph Projection/Orientation Classification: A Multi-institutional Study

**DOI:** 10.1101/2025.03.12.25322948

**Authors:** Yasuhito Mitsuyama, Hirotaka Takita, Shannon L Walston, Ko Watanabe, Shoya Ishimaru, Yukio Miki, Daiju Ueda

## Abstract

**Background:** Large-scale radiographic datasets often include errors in labels such as body-part or projection, which can undermine automated image analysis.

**Purpose:** To develop and externally validate two deep learning models—one for categorizing radiographs by body-part, and another for identifying projection and rotation of chest radiographs—using large, diverse datasets.

**Methods:** We retrospectively collected radiographs from multiple institutions and public repositories. For the first model (Xp-Bodypart-Checker), we included seven categories (Head, Neck, Chest, Incomplete Chest, Abdomen, Pelvis, Extremities). For the second model (CXp-Projection-Rotation-Checker), we classified chest radiographs by projection (anterior-posterior, posterior-anterior, lateral) and rotation (upright, inverted, left rotation, right rotation). Both models were trained, tuned, and internally tested on separate data, then externally tested on radiographs from different institutions. Model performance was assessed using overall accuracy (micro, macro, and weighted) as well as one-vs-all area under the receiver operating characteristic curve (AUC).

**Results:** In the Xp-Bodypart-Checker development phase, we included 429,341 radiographs obtained from Institutions A, B, and MURA. In the CXp-Projection-Rotation-Checker development phase, we included 463,728 chest radiographs from CheXpert, PadChest, and Institution A. The Xp-Bodypart-Checker achieved AUC values of 1.00 (99% CI: 1.00–1.00) for all classes other than Incomplete Chest, which had an AUC value of 0.99 (99% CI: 0.98– 1.00). The CXp-Projection-Rotation-Checker demonstrated AUC values of 1.00 (99% CI: 1.00–1.00) across all projection and rotation classifications.

**Conclusion:** These models help automatically verify image labels in large radiographic databases, improving quality control across multiple institutions.

## Introduction

Deep learning (DL) models using chest radiographs have made remarkable progress in recent years (1,2). We are now at a stage where more scalable and versatile models are being developed using larger, multi-institutional, and multinational datasets. Beyond disease detection, these models have evolved to accomplish tasks that are challenging for humans, such as estimating cardiac function, respiratory function, and biological age, offering substantial promise for improving diagnosis of chest radiograph (3–5). Consequently, there is a growing demand for more diverse datasets to power these advanced applications.

However, with the diversification and expansion of data, quality control for model training has emerged as a crucial challenge. As chest radiograph repositories increase in size and heterogeneity, ensuring correct labeling becomes more difficult, particularly because many images are stored in DICOM format, which relies on human entry for patient and examination information (6). In practice, errors and missing data frequently arise from manual or semi-automated workflows (7,8). Basic labels indicating the body-part and projection are critical both for clinical settings and data preprocessing (9). Inaccuracies in this metadata can undermine the reliability of subsequent DL-based analyses. These issues highlight the need for automated solutions that can detect mislabeled images before they negatively affect training and validation in deep learning pipelines.

To address these challenges, several machine learning and deep learning methods have been proposed for classifying radiographed body-parts, chest radiograph projection, or chest radiograph rotation (10–21). These automated systems aim to enhance standardization by catching potential metadata errors. However, many of these earlier studies relied on single-institution data or relatively small datasets, limiting their generalizability (11–19). Broader validation across multiple healthcare systems and regions is therefore vital to establish these models’ effectiveness in diverse clinical settings.

Against this background, we developed and externally tested two separate deep learning models using a large-scale, multi-institutional dataset. The first model, Xp-Bodypart-Checker, classifies radiographs of various body-parts, while the second model, CXp-Projection-Rotation-Checker, specializes in detecting projection and rotation of chest radiographs.

## Methods

### Study design

This multicenter retrospective study was conducted to develop and externally test two distinct DL models. The first model, Xp-Bodypart-Checker, classifies body-parts from various body-parts radiographic images. The second model, CXp-Projection-Rotation-Checker, determines both the projection (anterior-posterior [AP], posterior-anterior [PA], or lateral) and the rotation (upright, inverted, left rotation, or right rotation) of chest radiographs. The study followed the Declaration of Helsinki and was approved by the Institutional Review Board (approval numbers 2024-100, and 2022-0021K). The requirement for informed consent was waived by the ethics committees because only routinely acquired clinical images were used. This manuscript was prepared in compliance with the STARD guidelines (22).

### Participants, image collection, and examination acquisition

For Xp-Bodypart-Checker, radiographs of patients aged ≥18 years were retrospectively collected at Institution A (January 1, 2019, to December 31, 2021) and Institution B (January 1, 2023, to December 31, 2023). All radiographic examinations for each patient in this period were included. Additionally, upper extremity radiographs were collected from MURA (23). MURA is a large dataset of musculoskeletal radiographs containing 40,561 images from 14,863 studies. As age information was not available in MURA, all publicly available radiographs from this database were collected.

For CXp-Projection-Rotation-Checker, chest radiographs of patients aged ≥18 years were consecutively collected from Institution A (January 1, 2019, to December 31, 2021), as well as CheXpert (24) and PadChest (25). CheXpert is one of the largest publicly available chest radiograph datasets, comprising 224,316 chest radiographs from 65,401 patients who underwent radiographic examinations at Stanford Health Care between October 2002 and July 2017. The PadChest dataset is composed of 160,861 images from over 67,000 patients, collected from Hospital San Juan in Spain from 2009 to 2017. Since PadChest did not directly record patient age at examination, we excluded patients under 18 years old by calculating age from the ‘StudyDate_DICOM’ and ‘PatientBirth’ columns, and also excluded any pediatric protocols, corrupted images, or unknown birth years. If a patient had multiple, inconsistent gender entries, we labeled gender as unknown. All radiographic examinations for each patient in this period were included.

### Reference standard

For Xp-Bodypart-Checker, initially, all radiographs from Institutions A and B were classified into six categories (Head, Neck, Chest, Abdomen, Pelvis, and Extremities) based on the BodyPartExamined tag in their DICOM metadata, then verified and corrected by two board-certified radiologists (5 and 9 years of experience). To further refine the classification for images labeled as “Chest,” we examined whether both lung fields were fully captured or partly excluded, thereby subdividing this category into “Chest,” in which both lung fields were fully visible, and “Incomplete Chest,” in which one or both lung fields were partially excluded. Additionally, all radiographs from MURA were labeled as “Extremities”. As a result, the final classification encompassed seven distinct categories in total.

For CXp-Projection-Rotation-Checker, chest radiographs from Institution A were classified into three projections (AP, PA, and Lateral) based on the BodyPartExamined tag, ProtocolName tag, SeriesDescription tag, and StudyDescription tag in their DICOM metadata, then verified and corrected by two board-certified radiologists (5 and 9 years of experience). Using the imaging data recorded in each database (CheXpert and PadChest), we classified the chest radiographs according to their projection. Furthermore, rotation labels were randomly assigned to all chest radiographs to achieve an equal distribution (1:1:1:1 ratio) among four rotations: upright, inverted, left rotation, and right rotation. The images were then rotated according to these assigned rotation labels.

### Data partition

For Xp-Bodypart-Checker, radiographs from Institution A were randomly allocated on a per-patient basis in an 8:1:1 ratio for training, tuning, and internal test datasets, respectively. All radiographs from Institution B and MURA were utilized as the external test dataset.

For CXp-Projection-Rotation-Checker, chest radiographs from CheXpert and PadChest were randomly allocated on a per-patient basis in an 8:1:1 ratio for training, tuning, and internal test datasets, respectively. All chest radiographs from Institution A were utilized as the external test dataset.

### Model development

We developed two DL models using EfficientNet (26): one for classifying radiograph body-parts (Xp-Bodypart-Checker) and another for classifying projection and rotation of chest radiographs (CXp-Projection-Rotation-Checker). Cross-entropy loss was used as the loss function for both models. Each DL model was trained and tuned using the training and tuning datasets, respectively, starting with the initial parameters from the ImageNet-pretrained model and allowing the overall parameters to be updated. The model that achieved the smallest loss function value on the tuning dataset (within 100 epochs) was selected as the best-performing model. The longest side of each image was downscaled to 256 pixels while maintaining the aspect ratio, and the width along the shorter side was then padded black to 256 pixels. We performed data expansion with TrivialAugment Wide (27). Every development process was performed using PyTorch framework (version 2.0.1). Detailed processes for the development of the DL models and machine environment are shown in the appendix (p 2). Outlines of each of the two models are shown in Figures 1 and 2.

**Figure 1:**
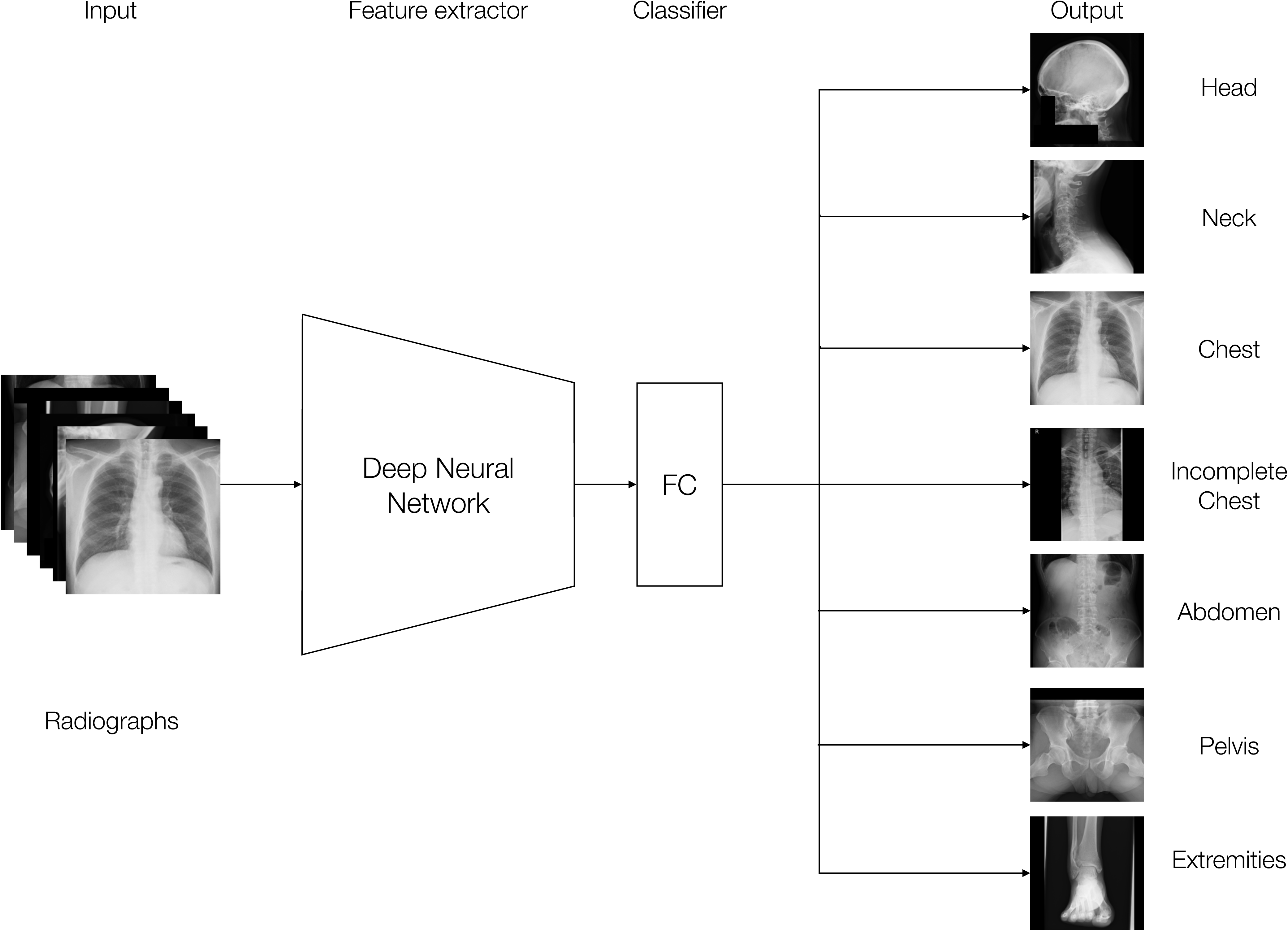
Outline of Xp-Bodypart-Checker The input various body-part radiograph is fed into an EfficientNet-based feature extractor, which progressively down-samples the image through multiple convolutional layers to learn high-level features. The final extracted feature map is then passed into a fully connected (FC) layer that serves as the classifier. This FC layer outputs one of seven body-part categories for the input radiographs.

**Figure 2:**
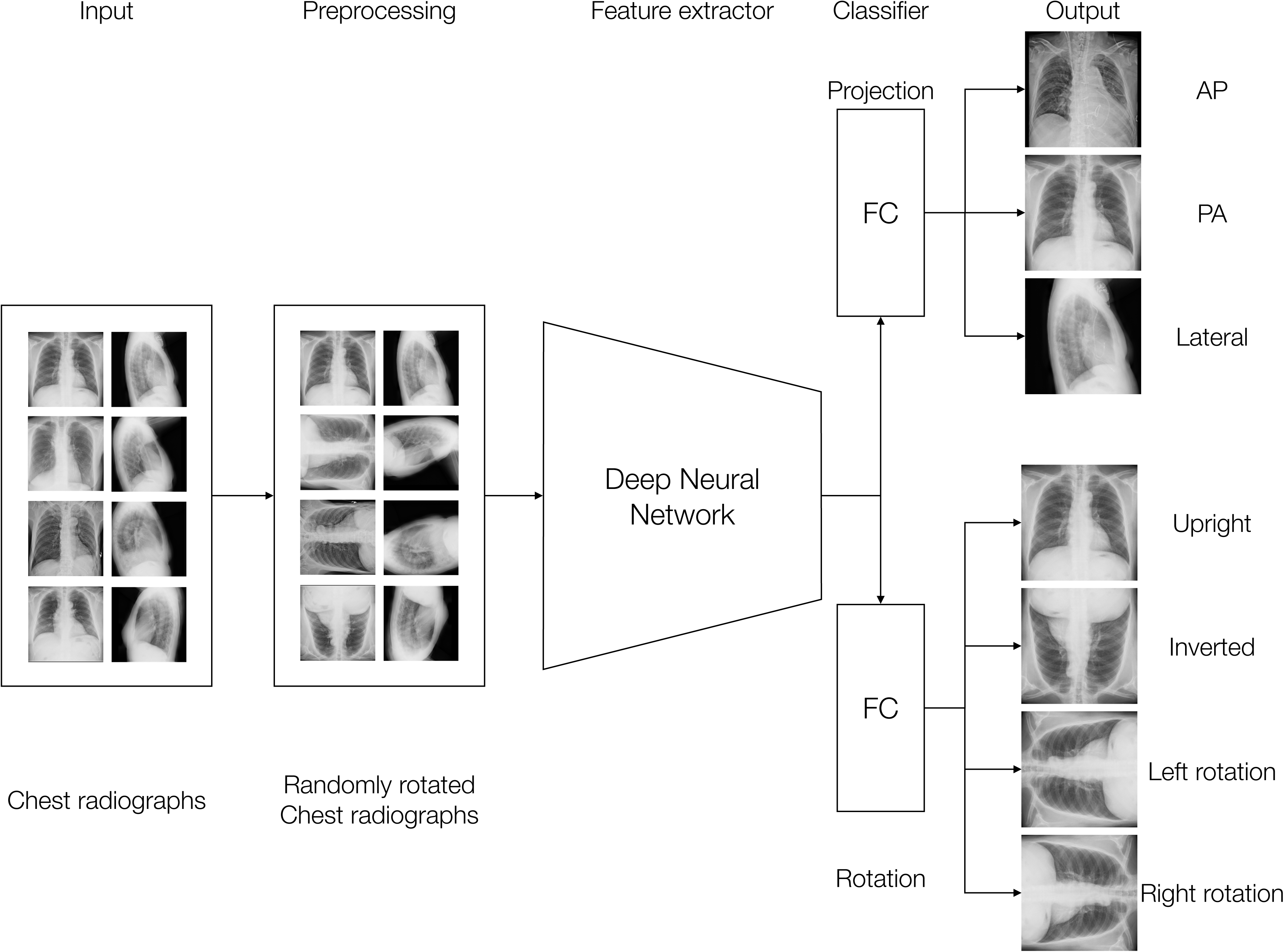
Outline of CXp-Projection-Rotation-Checker The input chest radiograph is fed into an EfficientNet backbone, which progressively down-samples and extracts hierarchical image features. The resulting feature representation is then passed into a fully connected (FC) layer configured to produce two classification outputs: one for chest radiograph projection (eg, anteroposterior, posteroanterior, or lateral) and another for chest radiograph rotation (eg, upright, inverted, left rotation, or right rotation). This multi-output design allows the model to learn both tasks simultaneously from a shared set of features.

### Model test

For Xp-Bodypart-Checker, we evaluated the classification performance of the DL model using the best model identified by the tuning dataset. The accuracy of body-part classification from radiographs was evaluated using an internal test dataset (Institution A) and external test datasets (Institution B and MURA).

For CXp-Projection-Rotation-Checker, we evaluated the classification performance of the DL model using the best model identified by the tuning dataset. The accuracy of projection classification and rotation classification from chest radiographs was evaluated using internal test datasets (CheXpert and PadChest) and an external test dataset (Institution A).

### Statistical analysis

For Xp-Bodypart-Checker, we calculated the overall model accuracy using micro-average, macro-average, and weighted-average. Furthermore, we calculated the accuracy for each body-part class and generated receiver operating characteristic (ROC) curves and area under the receiver operating characteristic curve (AUC) values. We created confusion matrices for the external test datasets (Institution B and MURA).

For CXp-Projection-Rotation-Checker, we calculated the overall model accuracy using micro-average, macro-average, and weighted-average for both projection and rotation classification tasks. Additionally, we calculated the accuracy for each projection class and rotation class, and generated ROC curves and AUC values. We created confusion matrices for the external test dataset (Institution A) for both projection and rotation classification tasks.

All analyses were done in SciPy using Python (version 3.8.1). These statistical inferences were performed with a two-sided significance level of 1%, and the performance metrics were presented with 99% confidence intervals.

## Results

### Datasets

For Xp-Bodypart-Checker, we included 297,846 radiographs obtained from 44,772 patients at Institution A, 91,490 radiographs from 12,895 patients at Institution B, and 40,005 radiographs from MURA. Details of these datasets are shown in Table 1, and the eligibility flowcharts are illustrated in the appendix (p 3). After review by two radiologists, the body-part labels based on the DICOM tags were modified for 3.9% of the radiographs from Institution A and 12.8% of those from Institution B, as summarized in the appendix (p 4).

**Table 1:** Demographics of datasets for Xp-Bodypart-Checker.

|  | Institution A |  |  | Institution B | MURA |
| --- | --- | --- | --- | --- | --- |
|  | Training | Tuning | Internal test | External test | External test |
| Total radiographs | 237475 | 31039 | 29332 | 91490 | 40005 |
| Total patients | 33388 | 5715 | 5669 | 12895 | N/A |
| Male | 15376 | 2575 | 2508 | 5308 | N/A |
| Female | 18012 | 3140 | 3161 | 7587 | N/A |
| Age (years) |  |  |  |  |  |
| Mean (±sd) | 62±17.2 | 63±16.7 | 63±16.7 | 71±14.3 | N/A |
| Range | 18–105 | 18–101 | 18–98 | 18–102 | N/A |
| Body-part shown in radiograph |  |  |  |  |  |
| Head | 1997 | 268 | 235 | 587 | 0 |
| Neck | 15623 | 1829 | 1952 | 2652 | 0 |
| Chest | 69560 | 9221 | 8813 | 26370 | 0 |
| Incomplete chest | 7797 | 1058 | 1058 | 655 | 0 |
| Abdomen | 68018 | 8645 | 8164 | 12468 | 0 |
| Pelvis | 17417 | 2443 | 1927 | 11613 | 0 |
| Extremities | 57063 | 7575 | 7183 | 37145 | 40005 |

Data are no. unless otherwise noted. sd denotes standard deviation.

For CXp-Projection-Rotation-Checker, we included 223,645 chest radiographs from 64,736 patients in CheXpert after excluding 3 chest radiographs for not meeting the age criterion. We included 152,573 chest radiographs from 62,935 patients in PadChest, with 7639 chest radiographs excluded. The majority of these (7,613 chest radiographs) were excluded for not meeting the age criterion, including 7,607 radiographs for which the examination-year minus the patient’s birth-year was <19, and 6 radiographs for which the birth year was unknown. Another 22 chest radiographs were excluded due to image corruption, and 4 chest radiographs excluded because they were acquired under a pediatric protocol. We included 87,510 chest radiographs from 25,228 patients at Institution A, after excluding 84 with undeterminable projection. Details of these datasets are shown in Table 2, and the eligibility flowcharts are illustrated in the appendix (p 5). After review by two radiologists, the projection labels based on the DICOM tags were modified for 3.3% of the chest radiographs from Institution A, as summarized in the appendix (p 6).

**Table 2:** Demographics of datasets for CXp-Projection-Rotation-Checker.

|  | CheXpert |  |  | PadChest |  |  | Institution A |
| --- | --- | --- | --- | --- | --- | --- | --- |
|  | Training | Tuning | Internal test | Training | Tuning | Internal test | External test |
| Total | 180002 | 21894 | 21749 | 121910 | 15509 | 15154 | 87510 |
| Total patients | 51979 | 6398 | 6359 | 50328 | 6376 | 6231 | 25228 |
| Male | 28837 | 3564 | 3516 | 24146 | 3020 | 2964 | 12693 |
| Female | 23142 | 2834 | 2843 | 26182 | 3356 | 3267 | 12535 |
| Unknown | 1 | 0 | 0 | 9 | 0 | 0 | 0 |
| Age (years) |  |  |  |  |  |  |  |
| Mean (±sd) | 60±18.6 | 60±18.5 | 60±18.4 | 59±17.5 | 59±17.5 | 59±17.6 | 63±16.8 |
| Range | 18–90 | 18–90 | 18–90 | 19–105 | 19–100 | 19–104 | 18–101 |
| Projection |  |  |  |  |  |  |  |
| Frontal | 154077 | 18739 | 18393 | 83743 | 10657 | 10406 | 78184 |
| AP | 130521 | 15900 | 15335 | 13444 | 1903 | 1626 | 27803 |
| PA | 23556 | 2839 | 3058 | 70299 | 8754 | 8780 | 50381 |
| Lateral | 25925 | 3155 | 3356 | 38167 | 4852 | 4748 | 9326 |

Data are no. unless otherwise noted. sd denotes standard deviation. AP = Anterior-Posterior, PA = Posterior-Anterior.

### Model evaluation

For Xp-Bodypart-Checker, the best-performing model was obtained at 11 epochs with the lowest loss value. The overall model performance was assessed through micro-, macro-, and weighted-accuracy calculations. For the external test set from Institution B, these accuracy metrics were 98.5% (99% CI: 98.4%–98.6%), 99.6% (99% CI: 99.6%–99.6%), and 99.4% (99% CI: 99.4%–99.5%), respectively. Additionally, we computed class-specific accuracy scores for each category; for example, the accuracy for the Chest class was 99.8% (99% CI: 99.7%–99.8%). To assess the model’s discriminative capability, we generated one-vs-all ROC curves for each class and calculated the corresponding AUC values, which ranged from 0.99 (99% CI: 0.98–1.00) for the Incomplete Chest class to 1.00 (99% CI: 1.00–1.00) for the Head, Neck, Chest, Abdomen, Pelvis, and Extremities classes. The ROC curves are presented in Figure 3A, and the accuracy and AUC values for both the overall performance and each category are summarized in Table 3. Confusion matrices were created to illustrate performance across all categories (Figure 3B). For the external test set from MURA, the micro-accuracy was 98.6% (99% CI: 98.4%–98.7%). The accuracy and AUC values for both the overall performance and each category are summarized in Table 3. Confusion matrices were created to illustrate performance across all categories (Figure 3C).

**Figure 3:**
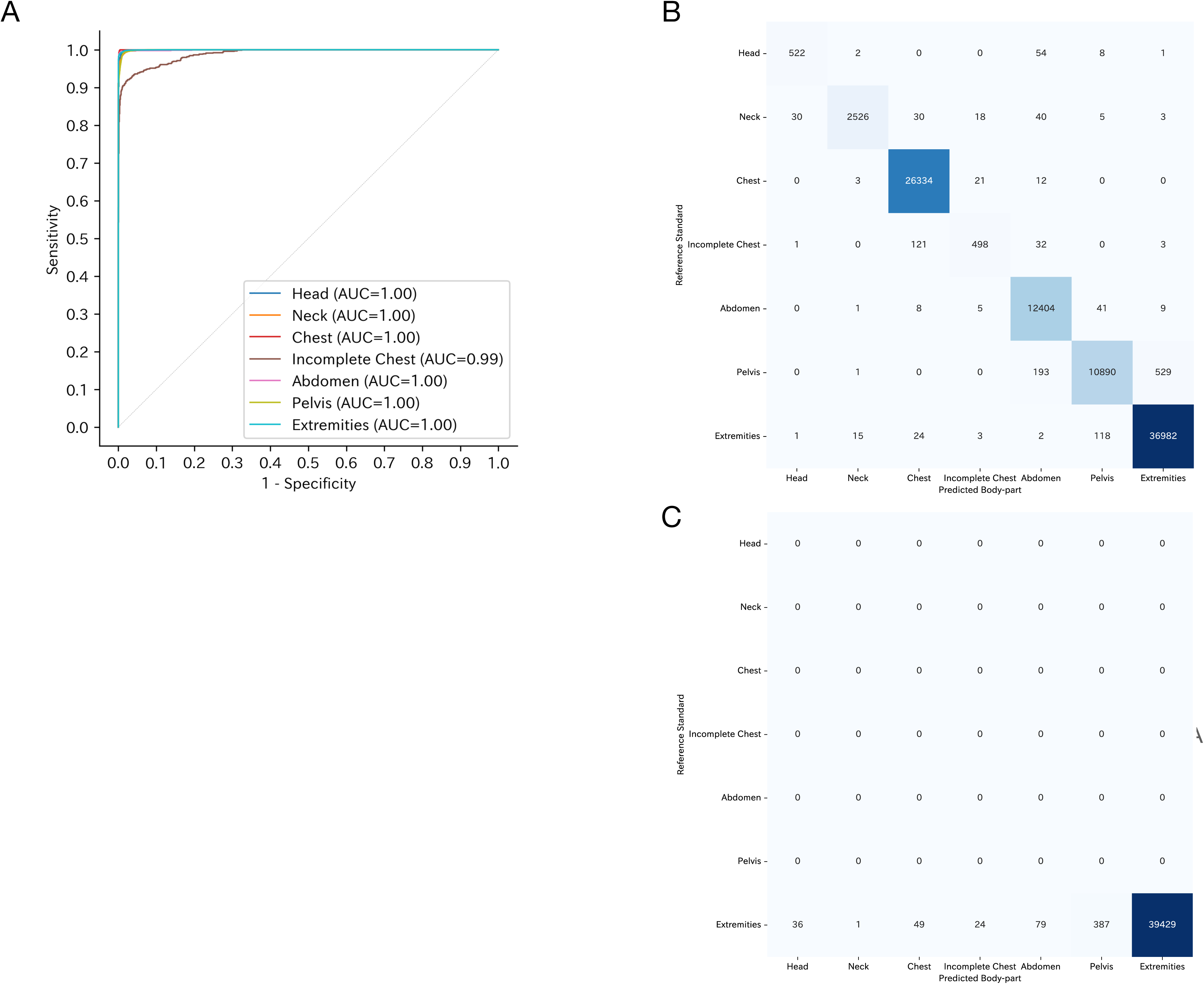
Receiver operating characteristic curves and confusion matrices of Xp-Bodypart-Checker (A) Receiver operating characteristic curves for body-part classification (seven categories) using the external dataset from Institution B. (B) Confusion matrix for body-part classification in the external dataset from Institution B. (C) Confusion matrix for body-part classification in the external dataset from MURA. For the external dataset from MURA, which contains only “Extremities” radiographs, no receiver operating characteristic curve is shown because there is only one class. AUC=area under the receiver operating characteristic curve.

**Table 3:** Results of Xp-Bodypart-Checker.

|  | Institution A |  | Institution B |  | MURA |  |
| --- | --- | --- | --- | --- | --- | --- |
|  | Internal test |  | External test |  | External test |  |
| Overall |  |  |  |  |  |  |
| Micro average accuracy (%) | 99.7 | (99.6–99.8) | 98.5 | (98.4–98.6) | 98.6 | (98.4–98.7) |
| Macro average accuracy (%) | 99.9 | (99.6–99.8) | 99.6 | (99.6–99.6) | N/A | N/A |
| Weighted average accuracy (%) | 99.9 | (99.6–99.8) | 99.4 | (99.4–99.5) | N/A | N/A |
| Body-part within radiographs |  |  |  |  |  |  |
| Head |  |  |  |  |  |  |
| Accuracy (%) | >99.9 | (99.9–100.0) | 99.9 | (99.9–99.9) | N/A | N/A |
| AUC | 1.00 | (1.00–1.00) | 1.00 | (1.00–1.00) | N/A | N/A |
| Neck |  |  |  |  |  |  |
| Accuracy (%) | >99.9 | (99.9–100.0) | 99.8 | (99.8–99.9) | N/A | N/A |
| AUC | 1.00 | (1.00–1.00) | 1.00 | (1.00–1.00) | N/A | N/A |
| Chest |  |  |  |  |  |  |
| Accuracy (%) | 99.8 | (99.8–99.9) | 99.8 | (99.7–99.8) | N/A | N/A |
| AUC | 1.00 | (1.00–1.00) | 1.00 | (1.00–1.00) | N/A | N/A |
| Incomplete chest |  |  |  |  |  |  |
| Accuracy (%) | 99.8 | (99.7–99.8) | 99.8 | (99.7–99.8) | N/A | N/A |
| AUC | 1.00 | (1.00–1.00) | .99 | (.98–1.00) | N/A | N/A |
| Abdomen |  |  |  |  |  |  |
| Accuracy (%) | 99.9 | (99.8–99.9) | 99.6 | (99.5–99.6) | N/A | N/A |
| AUC | 1.00 | (1.00–1.00) | 1.00 | (1.00–1.00) | N/A | N/A |
| Pelvis |  |  |  |  |  |  |
| Accuracy (%) | 99.9 | (99.9–100.0) | 99.0 | (98.9–99.1) | N/A | N/A |
| AUC | 1.00 | (1.00–1.00) | 1.00 | (1.00–1.00) | N/A | N/A |
| Extremities |  |  |  |  |  |  |
| Accuracy (%) | 99.9 | (99.9–100.0) | 99.2 | (99.2–99.3) | 98.6 | (98.4–98.7) |
| AUC | 1.00 | (1.00–1.00) | 1.00 | (1.00–1.00) | N/A | N/A |

Data are percentages (99% confidence interval) unless otherwise stated. AUC=area under the receiver operating characteristic curve.

For CXp-Projection-Rotation-Checker, the best-performing model was obtained at 3 epochs with the lowest loss value. We evaluated micro-, macro-, and weighted-accuracy for both projection and rotation classifications. On the external test set from Institution A, projection classification reached a micro-accuracy of 98.5% (99% CI: 98.3%–98.6%), macro-accuracy of 98.5% (99% CI: 98.3%–98.6%), and a weighted-accuracy of 98.5% (99% CI: 98.3%–98.6%). Rotation classification achieved a micro-accuracy of 99.3% (99% CI: 99.3%–99.4%), macro-accuracy of 99.3% (99% CI: 99.3–99.4%), and a weighted-accuracy of 99.3% (99% CI: 99.3%–99.4%). Additionally, we computed class-specific accuracy scores for each category within each classification. To assess the model’s discriminative capability, we generated ROC curves and calculated AUC values for each class using a one-vs-all approach. For projection classification, the AUC values were 1.00 (99% CI: 1.00–1.00) for the AP, PA, and Lateral view. For rotation classification, the AUC values were 1.00 (99% CI: 1.00–1.00) for the upright, inverted, left rotation, and right rotation view. The ROC curves are shown in Figure 4A and 4B, the confusion matrices are presented in Figure 4C and 4D, and the accuracy and AUC values both overall and for each category are presented in Table 4.

**Figure 4:**
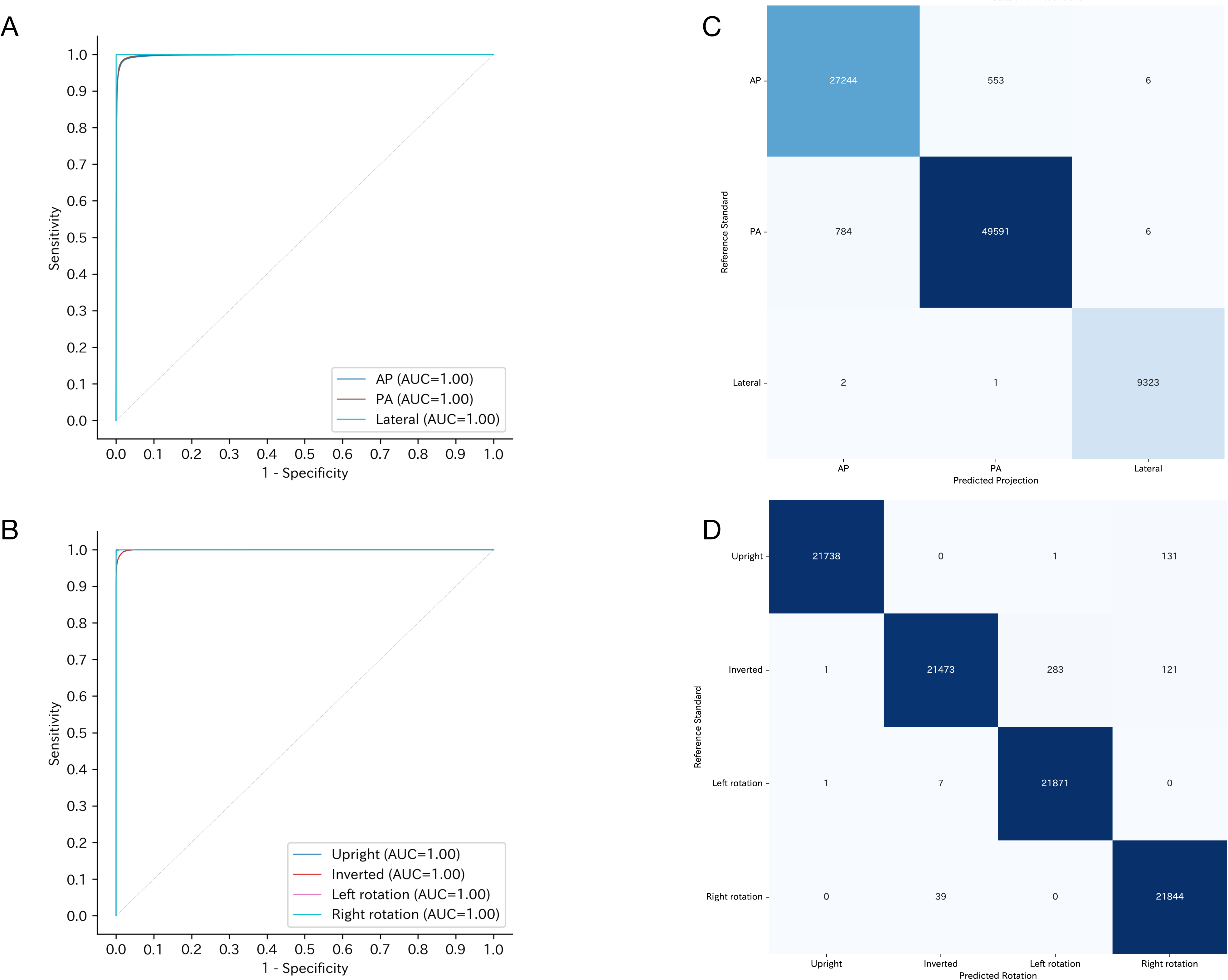
Receiver operating characteristic curves and confusion matrices of CXp-Projection-Rotation-Checker (A) One-vs-all receiver operating characteristic curves for projection classification (AP, PA, Lateral) using the external dataset from Institution A. (B) One-vs-all receiver operating characteristic curves for rotation classification (upright, inverted, left rotation, right rotation) using the external dataset from Institution A. (C) Confusion matrix for projection classification in the same external dataset. (D) Confusion matrix for rotation classification in the same external dataset. AP=Anterior-Posterior, PA=Posterior-Anterior. AUC=area under the receiver operating characteristic curve.

**Table 4:** Results of CXp-Projection-Rotation-Checker.

|  | CheXpert |  | PadChest |  | Institution A |  |
| --- | --- | --- | --- | --- | --- | --- |
|  | Internal test |  | Internal test |  | External test |  |
| Projection |  |  |  |  |  |  |
| Overall Accuracy |  |  |  |  |  |  |
| Micro average accuracy (%) | 99.4 | (99.3–99.5) | 91.2 | (90.8–91.7) | 98.5 | (98.3–98.6) |
| Macro average accuracy (%) | 99.4 | (99.3–99.5) | 91.2 | (90.8–91.7) | 98.5 | (98.3–98.6) |
| Weighted average accuracy (%) | 99.4 | (99.3–99.5) | 91.2 | (90.8–91.7) | 98.5 | (98.3–98.6) |
| Frontal |  |  |  |  |  |  |
| AP |  |  |  |  |  |  |
| Accuracy (%) | 99.4 | (99.3–99.5) | 96.8 | (96.5–97.1) | 98.5 | (98.4–98.6) |
| AUC | 1.00 | (1.00–1.00) | .98 | (.98–.98) | 1.00 | (1.00–1.00) |
| PA |  |  |  |  |  |  |
| Accuracy (%) | 99.4 | (99.3–99.5) | 92.4 | (91.9–92.8) | 98.5 | (98.4–98.6) |
| AUC | 1.00 | (1.00–1.00) | .99 | (.98–.99) | 1.00 | (1.00–1.00) |
| Lateral |  |  |  |  |  |  |
| Accuracy (%) | >99.9 | (99.9–100.0) | 93.4 | (93.0–93.8) | >99.9 | (99.9–100.0) |
| AUC | 1.00 | (1.00–1.00) | .99 | (.99–.99) | 1.00 | (1.00–1.00) |
| Rotation |  |  |  |  |  |  |
| Overall Accuracy |  |  |  |  |  |  |
| Macro average accuracy (%) | 99.9 | (99.8–99.9) | 94.3 | (93.9–94.7) | 99.3 | (99.3–99.4) |
| Micro average accuracy (%) | 99.9 | (99.8–99.9) | 94.3 | (93.9–94.7) | 99.3 | (99.3–99.4) |
| Weighted average accuracy (%) | 99.9 | (99.8–99.9) | 94.3 | (93.9–94.7) | 99.3 | (99.3–99.4) |
| Upright |  |  |  |  |  |  |
| Accuracy (%) | 99.9 | (99.9–1.00) | 97.1 | (96.9–97.4) | 99.8 | (99.8–99.9) |
| AUC | 1.00 | (1.00–1.00) | 1.00 | (1.00–1.00) | 1.00 | (1.00–1.00) |
| Inverted |  |  |  |  |  |  |
| Accuracy (%) | 99.9 | (99.9–1.00) | 97.2 | (96.9–97.5) | 99.5 | (99.4–99.5) |
| AUC | 1.00 | (1.00–1.00) | 1.00 | (1.00–1.00) | 1.00 | (1.00–1.00) |
| Left rotation |  |  |  |  |  |  |
| Accuracy (%) | 99.9 | (99.9–1.00) | 97.1 | (96.8–97.3) | 99.7 | (99.6–99.7) |
| AUC | 1.00 | (1.00–1.00) | 1.00 | (1.00–1.00) | 1.00 | (1.00–1.00) |
| Right rotation |  |  |  |  |  |  |
| Accuracy (%) | 99.9 | (99.9–1.00) | 97.3 | (97.0–97.5) | 99.7 | (99.6–99.7) |
| AUC | 1.00 | (1.00–1.00) | 1.00 | (1.00–1.00) | 1.00 | (1.00–1.00) |

Data are percentages (99% confidence interval) unless otherwise stated. AP=Anterior-Posterior, PA=Posterior-Anterior. AUC=area under the receiver operating characteristic curve.

In addition, to establish the characteristics of the misclassified images, we selected representative examples from the external test sets where the models’ predictions were incorrect. The example for Xp-Bodypart-Checker is shown in Figure 5A. These examples for CXp-Projection-Rotation-Checker are shown in Figure 5B, 5C and 5D.

**Figure 5:**
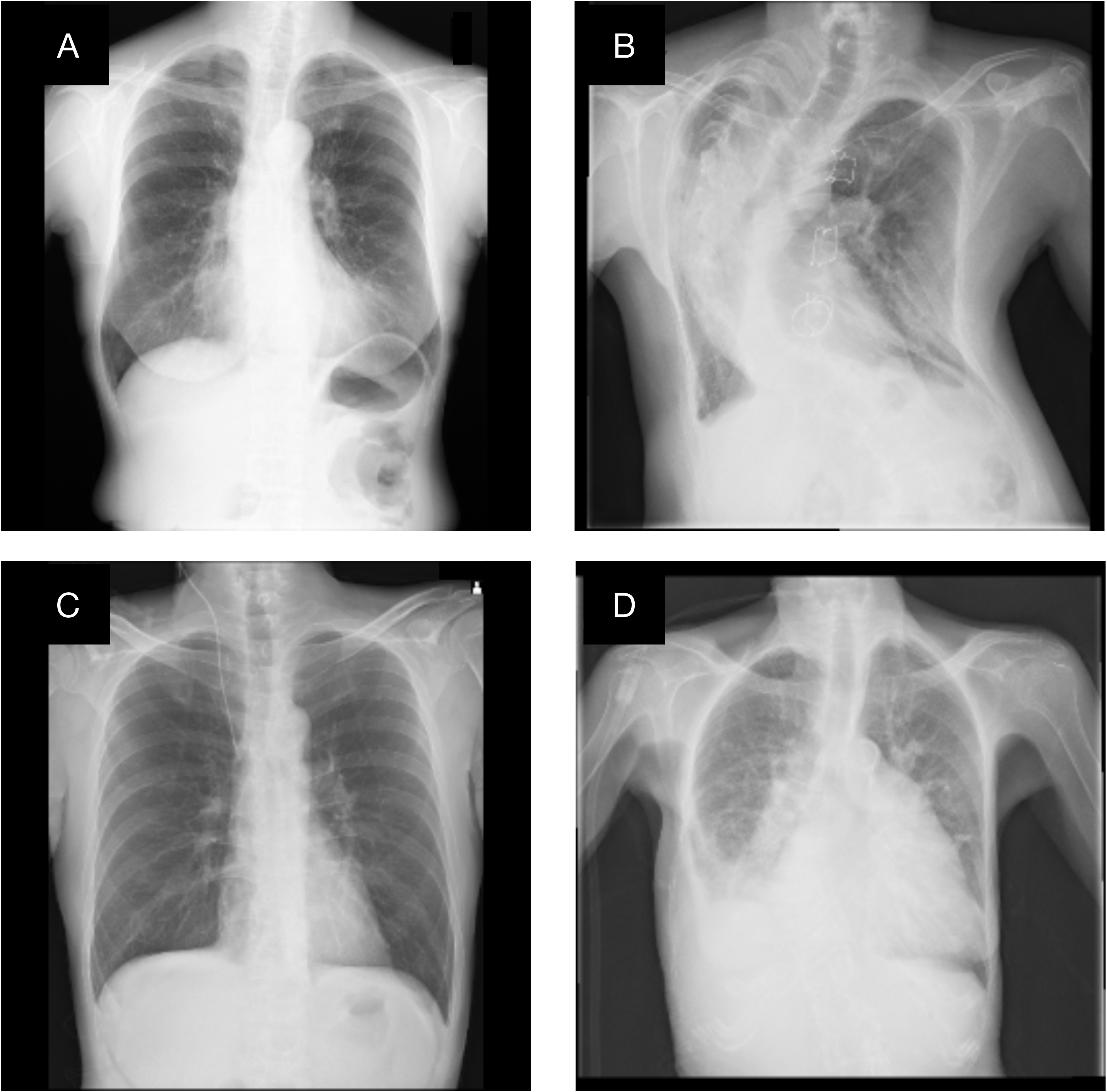
Representative examples of misclassifications by Xp-Bodypart-Checker and CXp-Projection-Rotation-Checker (A) Chest radiograph of a female in her 50s that was misclassified as “Abdomen” by Xp-Bodypart-Checker. (B) PA chest radiograph of a female in her 40s that was misclassified as “Lateral” by CXp-Projection-Rotation-Checker. (C) AP chest radiograph of a male in his 50s that was misclassified as “PA” by CXp-Projection-Rotation-Checker. (D) PA chest radiograph of a female in her 80s that was misclassified as “AP” by CXp-Projection-Rotation-Checker. AP=Anterior-Posterior, PA=Posterior-Anterior. AUC=area under the receiver operating characteristic curve.

## Discussion

In this study, we developed two deep learning models—Xp-Bodypart-Checker and CXp-Projection-Rotation-Checker—to address metadata inconsistencies in large-scale radiograph datasets. Xp-Bodypart-Checker was trained, tuned, and internally tested on data from Institution A and externally tested using data from Institution B and MURA. CXp-Projection-Rotation-Checker was trained, tuned, and internally tested using data from CheXpert and PadChest, then externally tested using data from Institution A. Altogether, over 400,000 radiographs were involved in developing Xp-Bodypart-Checker and over 460,000 chest radiographs were involved for CXp-Projection-Rotation-Checker. Xp-Bodypart-Checker classified radiographs into seven categories (Head, Neck, Chest, Incomplete Chest, Abdomen, Pelvis, and Extremities) with excellent performance, achieving AUC values higher than 0.99. Similarly, CXp-Projection-Rotation-Checker distinguished chest radiograph projection (AP, PA, and Lateral) and rotation (upright, inverted, left rotation, or right rotation) with AUC values of 1.00.

A main strength of our approach is that it tackles both various body-part radiograph classification and detailed chest projection/rotation classification using two separate models. Previous studies often addressed only one task—such as body-part recognition (10,12,19) or projection/rotation identification (13–18,20,21,28)— with smaller, or single-institution datasets. In contrast, we used data from multiple institutions with diverse imaging practices, which broadened our models’ applicability. In terms of performance, Xp-Bodypart-Checker achieved 98.5% accuracy on external data from Institution B, indicating that our results are comparable to—or potentially exceed—those of prior studies, which reported accuracies between 86% and 97.4% (10,12). The large scale of our dataset likely enhanced generalizability. Meanwhile, CXp-Projection-Rotation-Checker, trained on 463,728 chest radiographs from multiple countries, reached 98.5% accuracy for projection and 99.3% for rotation on external data from Institution A. Prior studies usually focused on either projection or rotation alone, with reported accuracies ranging from 96% to over 99% for projection and 88% to over 99% for rotation (13–18,28). Our findings suggest that combining both tasks in one network can meet or surpass these existing benchmarks while adapting to various clinical conditions.

Accurate DICOM labeling is important for both clinical settings and large-scale research endeavors. In our study, 3.9% of radiographs at Institution A and 12.8% at Institution B required body-part label corrections, while 3.3% of chest radiographs at Institution A had incorrect projection tags. In vast imaging archives, even minor error rates quickly add up to hundreds or thousands of mislabeled examinations. Clinically, such errors can delay patient care—for instance, mislabeling an “AP” projection as “PA” may affect orientation-based measurements (29). In research pipelines, incorrect labels can compromise DL training (30,31). By automating the detection of mislabeled studies, our models help ensure data integrity at the front end of both clinical workflows and research pipelines. This not only saves time and resources but also underpins safer clinical decisions and more trustworthy scientific findings.

Despite the high overall accuracy, we found interesting results and areas for improvement through error analysis. Xp-Bodypart-Checker sometimes mislabeled chest radiographs that included the upper abdomen as “Abdomen.” Overlapping structures, such as the diaphragm and lower thorax, can blur the boundary between the chest and abdomen, suggesting the need for more flexible labeling or continuous scores in borderline cases. Similarly, CXp-Projection-Rotation-Checker struggled with certain anatomical variations. We observed that chest radiographs with a noticeably large cardiac silhouette in PA views were occasionally misclassified as AP, while those with a smaller cardiac silhouette in AP views were sometimes mistaken for PA. This likely reflects the known tendency for AP images to exaggerate heart size (29). In addition, PA chest radiographs showing marked spinal scoliosis were at times confused with lateral views, implying that the model may heavily rely on spine orientation cues. These findings indicate that training on more diverse cases and incorporating additional clinical context—such as patient posture or spinal alignment—could further enhance reliability.

This study had several limitations. First, the study did not include certain populations (e.g., pediatric patients) or full-body scans that include both trunk and extremities, limiting immediate relevance to those settings. Second, institution-specific labeling protocols might introduce biases, and integrating these models into established systems could require technical adjustments.

Our two-model strategy provides a practical way to detect and fix common labeling errors in large radiographic collections. By automatically verifying body-part, projection and rotation tags, it lays a stronger groundwork for routine clinical tasks and research projects. Future efforts will involve real-time testing, applications in pediatric or more complex imaging scenarios, and possible multi-label outputs for borderline cases. We believe that robust, automated labeling is a key step toward more reliable and versatile radiographic databases.

## Data Availability

The Deep Learning models developed in this study are publicly available on Hugging Face (https://huggingface.co/spaces/MedicalAILabo/Xp-Bodypart-Checker, https://huggingface.co/spaces/MedicalAILabo/CXp-Projection-Rotation-Checker).
The code developed in this study is publicly available on GitHub (https://github.com/Medical-AI-Lab/Nervus/).
However, chest radiographs from Institutions A and B are not available at this time, as these have been withheld by the hospitals that participated in the trials to protect participant privacy.
Additionally, data from CheXpert, PadChest, and MURA were used for this study; these datasets are accessible through their respective repositories (see References 23, 24, and 25).

## Acknowledgement

We used ChatGPT based on the GPT-4 architecture (September 25 Version; OpenAI; https://chat.openai.com/) to proofread the English in the manuscript, and the output was reviewed and approved by the authors. We are grateful to the National Hospital Organization Osaka Minami Medical Center for providing the data for this study.

## Data sharing statement

The Deep Learning models developed in this study are publicly available on Hugging Face (https://huggingface.co/spaces/MedicalAILabo/Xp-Bodypart-Checker, https://huggingface.co/spaces/MedicalAILabo/CXp-Projection-Rotation-Checker).

The code developed in this study is publicly available on GitHub (https://github.com/Medical-AI-Lab/Nervus/).

However, chest radiographs from Institutions A and B are not available at this time, as these have been withheld by the hospitals that participated in the trials to protect participant privacy.

Additionally, data from CheXpert, PadChest, and MURA were used for this study; these datasets are accessible through their respective repositories (see References 23, 24, and 25).

