## Supplemental Materials for "Deep Learning Models for Radiography Body-part Classification and Chest Radiograph Projection/Orientation Classification: A Multi-institutional Study"

**Table of Contents:**

**Section S1: Supplementary Methods**

1) Detailed process for parameter tuning of the deep learning model

2) Machine environment

**Section S2: Supplementary Figures**

Appendix Figure 1: Eligibility flowchart for Xp-Bodypart-Checker

Appendix Figure 2: Changes in body-part label after two radiologists’ review

Appendix Figure 3: Eligibility flowchart for CXp-Projection-Rotation-Checker

Appendix Figure 4: Changes in projection label after two radiologists’ review

**Section S3: References for the Supplementary Appendix**

**Section S1: Methods**

**1) Detailed process for parameter tuning of the deep learning model**

In this study, we developed two deep learning models, Xp-Bodypart-Checker and CXp-Projection-Rotation-Checker. Both models were built on the EfficientNetB4 architecture. For each model, we performed hyperparameter tuning for the optimizer, learning rate, and batch size. Specifically, we employed Adam as the optimizer for both models (the learning rate was searched within 0.001–0.05) and a batch size of 32 was used.

**2) Machine environment**

We adopted Ubuntu 20.04 (Canonical, London, England) with the PyTorch deep learning framework (version 2.0.1; The Linux Foundation; https://pytorch.org), with CUDA 11.8 (Nvidia Corporation, Santa Clara, CA) dependencies for graphics processing unit acceleration. We used a GDEP Advance Deep Learning Box equipped with four NVIDIA Titan V graphics processing units (Nvidia Corporation).

**Section S2: Supplementary Figures**

**Appendix Figure 1: Eligibility flowchart of Xp-Bodypart-Checker**
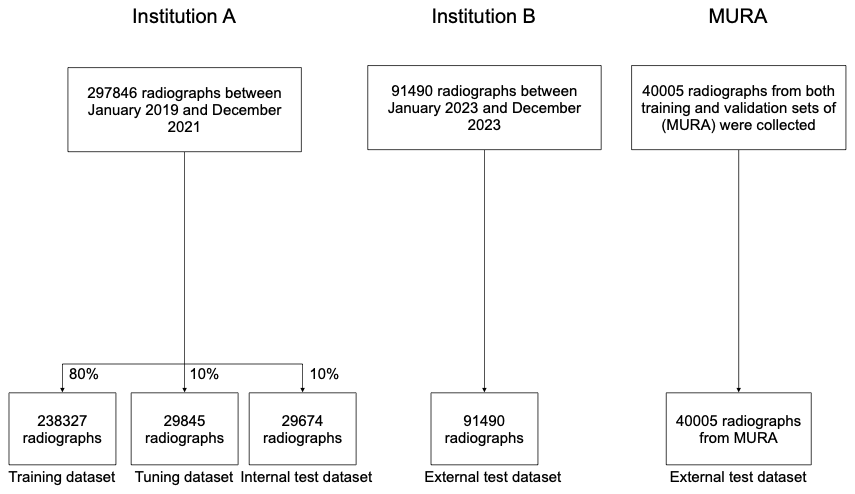
 These flowcharts describe the training, tuning, internal test, and external test datasets used during the development phase of Xp-Bodypart-Checker.

**Appendix Figure 2: Changes in body-part label before and after two radiologists’ review**

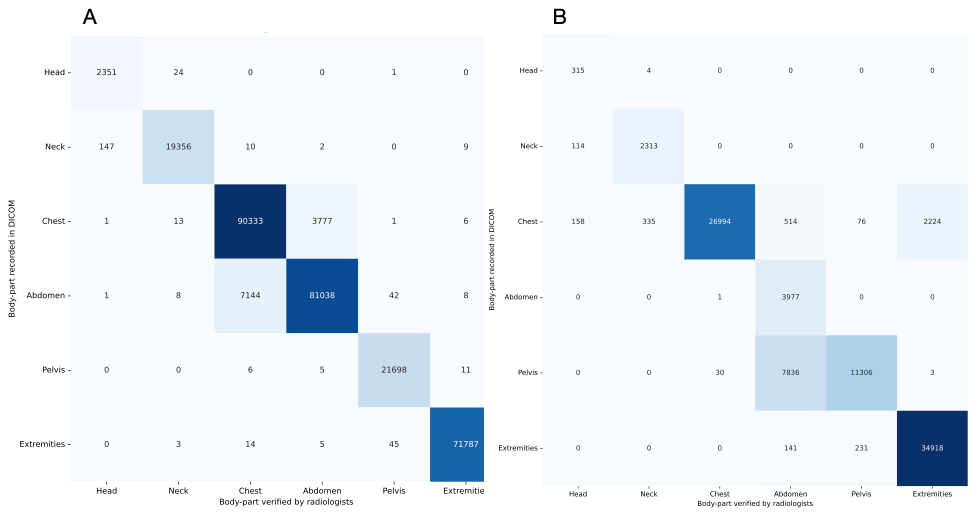

(A) Changes in body-part label of radiographs from Institution A before and after two radiologists’ review. (B) Changes in body-part label of radiographs from Institution B before and after two radiologists’ review.

The rows list the original body-part labels from the DICOM metadata, and the columns list the body-part labels assigned after verification by two board-certified radiologists. Each cell shows how many radiographs were confirmed or corrected. The diagonal cells where the row and column labels match represent the number of radiographs where the radiologists agreed with the original DICOM labels, while the off-diagonal cells show the number of radiographs that were changed.

**Appendix Figure 3: Eligibility flowchart of CXp-Projection-Rotation-Checker**

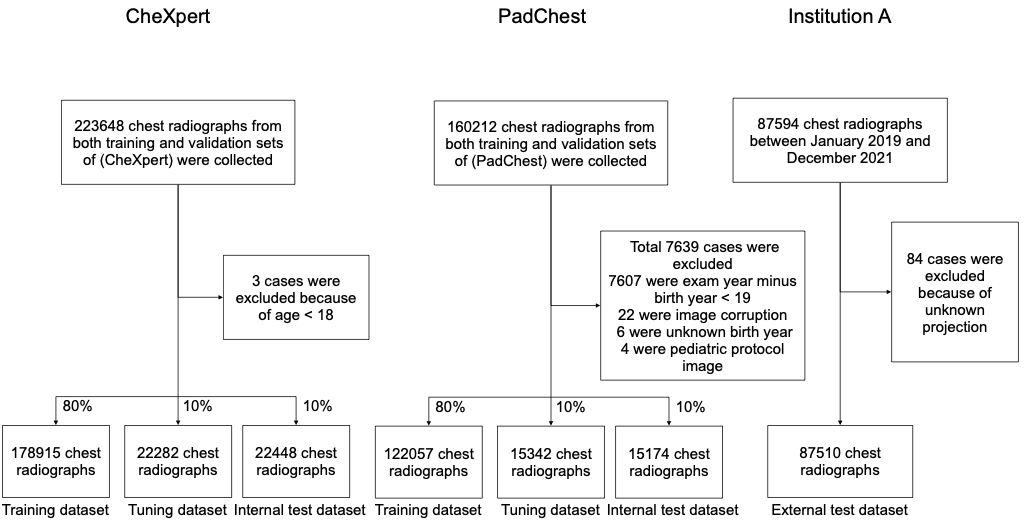
 These flowcharts describe the training, tuning, internal test, and external test datasets in the development phase of CXp-Projection-Rotation-Checker.

**Appendix Figure 4: Changes in projection label after two radiologists’ review**

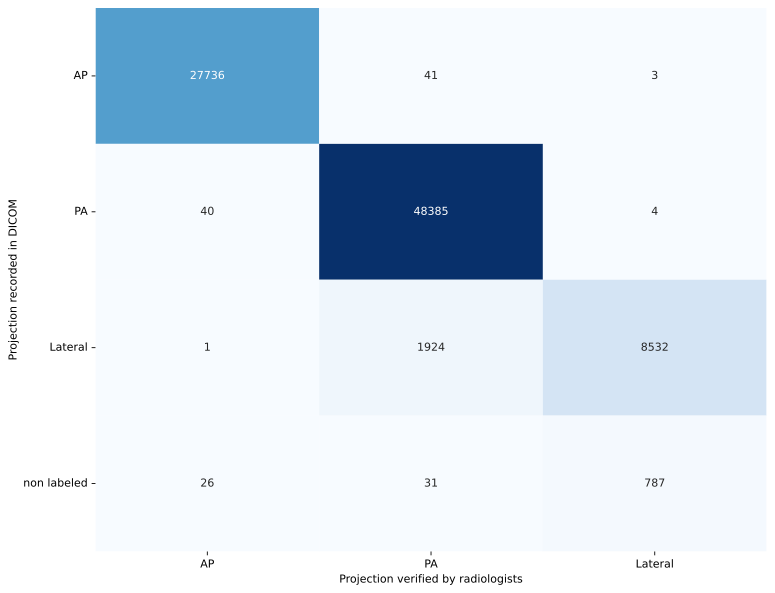

Changes in projection label of chest radiographs from Institution A before and after two radiologists’ review.

The rows list the original projection labels from the DICOM metadata, and the columns list the projection labels assigned after verification by two board-certified radiologists. Each cell shows how many chest radiographs were confirmed or corrected. The diagonal cells where the row and column labels match represent the number of chest radiographs where the radiologists agreed with the original DICOM labels, while the off-diagonal cells show the number of chest radiographs that were changed.
